# Comparing Providers for the Treatment of Early Childhood Caries with a Value-based Rating Methodology: A Cross-sectional study

**DOI:** 10.1101/2023.05.03.23289278

**Authors:** David Okuji, Betty Ben Dor, Ashiana Jivraj, Yinxiang Wu

## Abstract

**Purpose:** An adapted Porter-Teisberg model of value-based health outcomes and cost was applied to comparatively rate providers who treat children with early childhood caries, by treatment modalities, facilities, and geographic location.

**Methods:** Quality was measured by a Parental-Reported Symptom and Service Quality questionnaire, adapted from the Early Child Oral Health Impact Scale, Child-Oral Impacts on Daily Performance, and Dental Consumer Assessments of Healthcare Providers and Systems tools. Cost was measured by the labor cost for all providers involved in treatment and adjusted by dividing by total relative value units for the types and total number of procedures for all treatment visits. Value was calculated as the quotient of the weighted quality score and the adjusted cost, multiplied by a coefficient. To compare the value score by states and by facilities, Tukey’s multiple comparison test was used and a sensitivity analysis using the non-parametric Dunn’s Kruskal-Wallis multiple comparison test was conducted.

**Results:** For surgical treatment with general anesthesia, Tennessee ranked the highest, followed by Arizona, Hawaii, and New York. These four states had significantly higher value metrics than Florida, Massachusetts, and Maryland. Maryland had a significantly higher value score for surgical treatment without general anesthesia. While patients were seen for non-surgical treatment only in Maryland and New York, Maryland also had a significantly higher value for this treatment modality.

**Conclusion:** A value-based rating system of providers, by geographic region and treatment facility, can assist patients to better compare and select providers; third-party payers, such as insurance companies and government programs, to better compare outcomes and costs for provider systems and clinic facilities; and providers to benchmark their comparative ratings on an internal and external basis for continuous quality improvement.

**Highlights:**

- Non-surgical management of early childhood caries yields the most value.
- Surgical intervention with or without general anesthesia is less valued by patients.
- Ranking treatment modalities by value is a useful tool for patients.
- Using a parental-reported symptom and service quality questionnaire assesses quality.
- Provider value can be determined by dividing quality of care by the cost of delivery.

## INTRODUCTION

Recent years have seen an increased legislative and political effort to establish a value-based health care system in an attempt to deliver better care to patients while decreasing overall costs.^1^ A predominant model of evaluating and rating dental providers in the United States (**U.S**.) is through the use of online customer satisfaction and review digital media platforms, such as Yelp.com and Healthgrades.com.^2^ These public platforms allow patients to freely comment on their experiences with particular offices and providers in a manner that is easily accessible to potential and current patients, allowing them to use this information for the selection of a dentist. Customers who have positive experiences tend to submit high ratings and those who have negative experiences do not typically submit ratings, which can artificially inflate the overall ratings.^3^

One of the most comprehensive value-based care online platforms is the Medicare Care Compare tool, developed by the Centers for Medicare and Medicaid Services (**CMS**) and based on outcomes data.^4^ However, one large study showed Yelp.com was the most frequently used online surgeon rating site, and was nearly three times as likely to be used as the Medicare Physician Compare website.^2^ The Medicare Care Compare tool provides the public with a comprehensive website that patients can use to find in-network medical providers.^5^ Patients are provided information on the providers’ certifications and locations and have access to federal government provider-rating data. Few dentists are listed as providers, except participating oral and maxillofacial surgeons. Quality scores are listed as star ratings, are not typically available at the individual provider level, and are available at the “group affiliation” level if the individual provider and group opt to participate in the quality initiatives. The quality star ratings “are only publicly reported if the measured data meet the established public reporting standards and resonate with users.^6^” The quality star rating methodology is based upon establishing statistically derived benchmarks based upon mean performance across top-ranked performers.^6^ Participating providers who meet or exceed the benchmark are assigned 5 stars. An equal range method is utilized to assign 4-, 3-, 2-, and 1-star ratings.^6^ The benchmarks for providers include innovative model participation, electronic health record technology performance information, improvement activities, quality performance information, and/or patient survey scores.^7^

Neither customer satisfaction and review platforms nor the statistically derived CMS quality star ratings include the cost of delivering care as part of their rating methods. Hence, this opens an opportunity for an improved, standardized, evidence-based, and easily utilized approach for rating providers, including dentists, for the delivery of value-based health care.

Multiple organizations are studying evidence-based structure and processes toward value-based outcome models. The International Consortium for Health Outcomes Measurement (**ICHOM**)^8^ has based their approach on the 2006 book, *Redefining Health Care*,^9^ by Harvard Business School Professors Michael E. Porter and Elizabeth O. Teisberg, who respectively lead the Harvard Business School, Institute for Strategy & Competitiveness, Health Care Team and the Value Institute for Health and Care out of the University of Texas at Austin Dell Medical School and McCombs School of Business.^10,11^ Although it is quality-focused, not value-focused, the American Dental Association (**ADA**) has developed a Dental Quality Alliance (**DQA**) with the expressed mission of expanding a set of performance measures for the delivery of oral health care.^12^

Value-based studies which focus on early childhood caries (**ECC**) allow investigators to conduct research for a disease condition confined to a relatively short time-period, with known risk factors, and standard treatment modalities with wide variations in cost of care. ECC refers to caries found in the primary dentition of children under the age of six years old and globally presents as one of the most common chronic childhood diseases, primarily affecting those in low income and minority groups.^13^ It can lead to pain, infection, and decreased body mass index, along with negative effects on childhood learning and quality of life.^13,14^ ECC care presents a tremendous burden not only on families and communities, but on health care systems as well. As of 2013, pediatric oral health care generated $26.5 billion in expenditures.^15^ Given the negative clinical and economic impacts of ECC on the pediatric population, it makes financial sense to support patients and third-party payers with value-based tools to better select providers who improve oral health outcomes and reduce costs.

The Porter and Teisberg model for value-based health care focuses on the delivery of cycles of care for medical conditions.^9^ The model defines health care value as the quotient of quality and cost.^9^ The aim with this study is to use an adapted formulation of the Porter-Teisberg value-based model to rate provider systems for their management of ECC, with the specific objective to compare value scores and letter grade ratings of providers at the geographic location and clinic facility levels. The null hypothesis was that providers would demonstrate equal value scores and letter grade ratings, as determined by a value-based statistical model used in this study.

## METHODS

### Study Design and Setting

This study utilized an analytical, observational, cross-sectional design to compare a value score and letter grades for providers at the levels of treatment facilities and geographic regions identified by specific U.S. states. The study was approved by the NYU Grossman School of Medicine Institutional Review Board under protocol number 16-00331.

A convenience sampling methodology was utilized. Children under six years old, who were patients of federally qualified health centers (**FQHCs**) with a diagnosis of ECC, were recruited and stratified by the treatment modality selected by their legal caregiver via the informed consent process. The three treatment modalities included minimally invasive non-surgical care (**NSC**), surgical restorative care without adjunctive sedation or general anesthesia (**SC**), and surgical restorative care with general anesthesia (**SCGA**).

Value of care was calculated as the parental-reported quality divided by the total cost of care. Quality was measured by a Parental-Reported Symptom and Service quality Questionnaire (**PRSSQ**), which was completed by the caregiver approximately one-month after the last treatment visit (Figure 1). Total cost of care was calculated by multiplying the total number of hours to complete the treatment by the total hourly labor costs of health care providers (**HCP**) who were directly involved with the treatment. Value was then calculated by dividing the quality PRSSQ score by the total cost of care. The comparative value for each treatment facility and each region by state were statistically clustered and assigned a letter score, with “A” as the highest and “E” as the lowest ratings.

**Figure 1.**
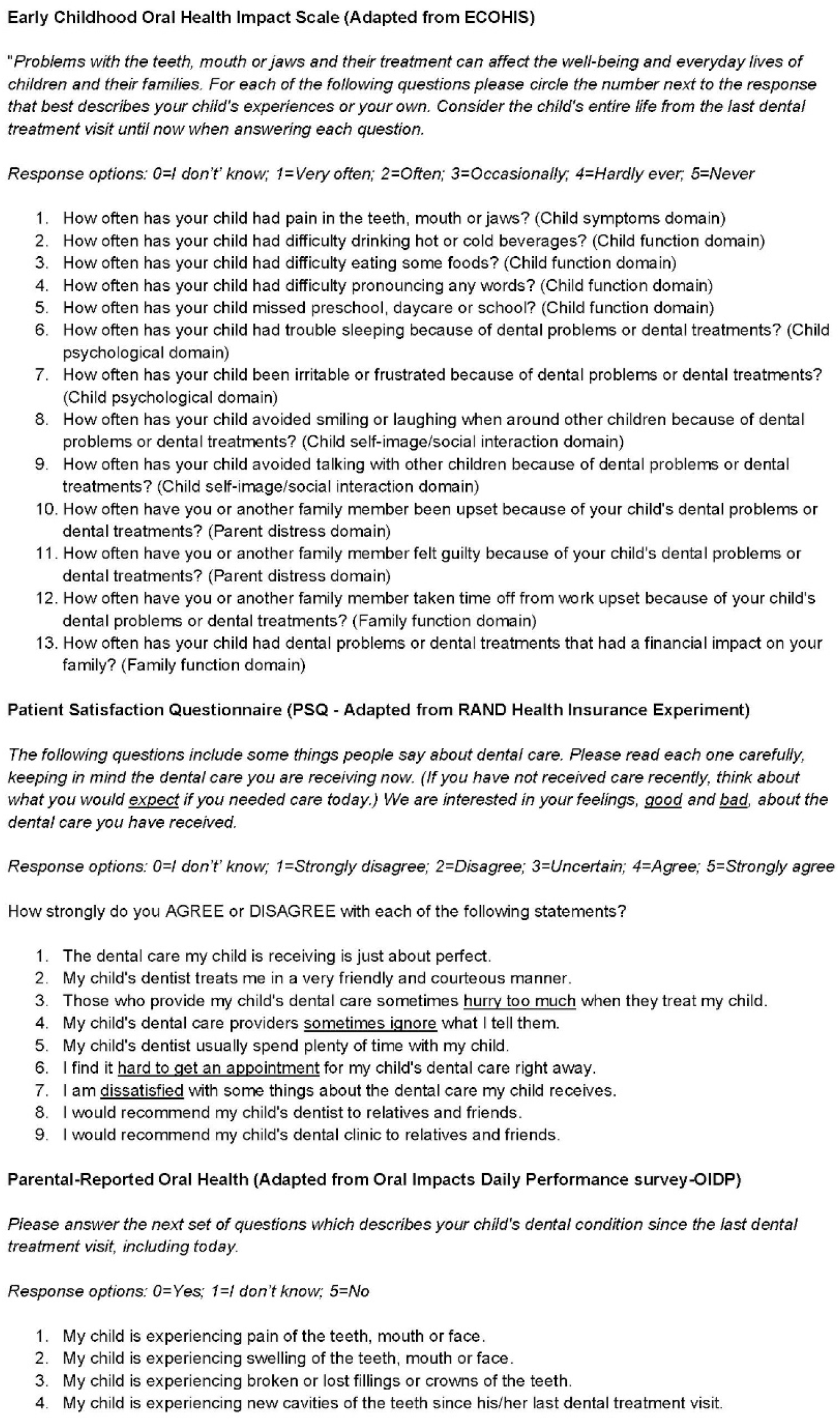
Post-treatment Parental-Reported Symptom and Service quality Questionnaire (PRSSQ), 26-questions, English version.

### Study Size and Participants

Patient care was carried out at 16 clinic facilities located at FQHCs, hospitals, and surgery centers affiliated with the NYU Langone Dental-Advanced Education in Pediatric Dentistry program in seven states: Arizona, Florida, Hawaii, Maryland, Massachusetts, New York, and Tennessee.

Four-hundred, eighty-seven (487) subjects were recruited from 12/01/2015 to 10/03/2016. The patient populations served at these health centers were from predominantly underserved communities and made up an ethnically diverse group. Parents or legal caregivers of pediatric patients with a diagnosis of ECC were recruited for the study and underwent the informed consent process if they assented to participation.

The post-hoc power analysis was calculated at 99.4%, based upon the study’s sample size, treatment size effects, and alpha at 0.05.

Inclusion criteria for the study were 1) registration as a patient at one of the FQHCs, hospitals, or surgery centers affiliated with the NYU Langone Dental-Advanced Education in Pediatric Dentistry program, 2) age six or younger at the time of enrollment into the study, 3) a diagnosis of ECC, 4) a full complement of 20 primary teeth, 5) American Society of Anesthesiology status of 1, 2, or 3, and 6) consent for treatment by the parent/caregiver in either Spanish or English.

### Variables

The independent variables for treatment modality were defined as follows:

**Non-surgical care (NSC)**: Non-surgical care was defined as treatment with topical fluoride varnish and silver diamine fluoride (**SDF**) applications, interim therapeutic restorations, motivational interviewing, frequent follow-up treatment visits, and active surveillance visits.

**Surgical care without sedation/general anesthesia (SC)**: Surgical care without sedation or GA was defined as full- or partial-mouth treatment with local anesthesia in one or more treatment visits with or without the use of nitrous oxide analgesia.

**Surgical care with general anesthesia (SCGA)**: Surgical care with general anesthesia (**GA**) was defined as full-mouth dental rehabilitation in one visit under GA.

The dependent variables for quality and cost were defined as follows:

**Quality (Q)**: Quality of care was assessed using a 26-question PRSSQ. This post-treatment survey was created for deployment to the parents or legal guardians of the patients one month after completion of treatment. The PRSSQ consists of questions adapted and compiled from three validated satisfaction surveys. Thirteen questions came from the Early Child Oral Health Impact Scale (**ECOHIS**),^16^ which looks at the effects of pediatric oral health on children and caregivers. Another six questions came from the Child-Oral Impacts on Daily Performance (**C-OIDP**),^17^ which assesses the impact of oral health on eating, speaking, cleaning teeth, smiling, emotional stability, relaxing, doing schoolwork, and social contact. Nine questions were sourced from the Dental Consumer Assessments of Healthcare Providers and Systems (**D-CAHPS**),^18^ which is used to measure satisfaction with dental care among consumers. The score for quality was calculated by using the equally weighted average score of the three components that made up the PRSSQ. Thus, as a metric, the PRSSQ is a compilation of existing, validated assessments for parental satisfaction, parent clinical observations, and impact on families.

**Cost (C)**: As a proxy for total direct cost, as prescribed by the Porter-Teisberg model, total direct labor costs associated with treatment were utilized since direct labor cost contributes to nearly 60% of operational expenses for a pediatric dental practice.^19^ Accountants define “total direct cost” as the cost of materials and supplies, in addition to labor. This study defined “total direct labor cost” as the total labor cost for HCPs required to complete the treatment of each child. The HCPs involved in delivering care included dental residents, attending dentists, anesthesiologists, nurse-anesthetists, nurses, surgery technicians, dental hygienists, and/or dental assistants. Direct labor cost was measured, and adjusted for the effects of the types and number of total procedures, by calculating the total duration of treatment time for the total number of visits, multiplying by the labor rate for the aggregate of each clinical staff member involved with the child’s treatment, and dividing by the total Relative Value Units (**RVU**) for the types and total number of procedures performed for all treatment visits.^20^ In this way, the RVU-adjusted cost provided a basis for equal comparison among the three treatment modalities. **Value (V)**: Value was calculated as quality (PRSSQ score), divided by RVU-adjusted cost (**C**), times a coefficient of 100, with the multiplier product resulting in a value numeric which is greater than zero.

**Letter grade ratings:** Based upon the statistical model, letter grade ratings were assigned to provider groups which were clustered to have statistically similar mean value scores. The highest assigned letter grade was (A) and the lowest (E).

The dependent variables for provider levels were defined as follows:

**Geographic location**: The location was defined as the U.S. state in which the treatment facilities are located and identified as Arizona (**AZ**), Hawaii (**HI**), Florida (**FL**), Maryland (**MD**), Massachusetts (**MA**), New York (**NY**), and Tennessee (**TN**).

**Treatment facility**: The 16 treatment facilities were classified by type of clinic facility, geographic location, and identified as 1) Out-patient dental clinics: AZ-1, FL-1, FL-2, MA-1, MD-1, NY-1, TN-1, 2) Out-patient dental clinics with mobile general anesthesiology services: AZ-1, NY-1, and TN-1, and 3) Hospital or ambulatory surgery centers: HI-1, HI-2, FL-3, FL-4, MA-2, MD-2, MD-3, NY-2, TN-2.

**Individual dentist-provider**: Although data was available at the individual dentist-provider level, each individual dentist-provider did not treat a sufficiently large number of patients to yield meaningful and comparative statistical analyses.

### Data Sources, Measurements, and Data Collection

#### Dental record

Patients first received an oral examination, with a diagnosis of ECC as defined by the American Academy of Pediatric Dentistry,^21^ followed by development of treatment plan options to be provided to the parent/caregiver. The children were then classified as an American Society of Anesthesiologists Physical **(ASA)** Status Classification^22^ 1, 2, or 3 and were categorized to receive either non-surgical care (**NSC**), surgical treatment without general anesthesia (**SC**), or surgical treatment with general anesthesia (**SCGA**).

Patients were treated by the assigned individual dentist-provider on any given day. Given the large number of dentist-providers rendering the treatments across all patients, the assumption was there should be very little variation in the treatment outcomes on average. Value was not able to be compared at the individual dentist-provider level because there was, on average, a very low number of treatments rendered per each dentist-provider. Individual providers were therefore grouped by state and clinic facilities. The maximum number of treatment visits for each treatment modality listed was one for SCGA and up to five visits for SC and NSC.

#### Third party sources

Direct labor cost of care was calculated by multiplying the duration of treatment time in hours by the aggregate hourly wage rate for HCPs involved in the treatment. The duration of treatment time was measured from the time of ingress to time of egress from the treatment room. The average hourly wage rate for HCPs was calculate from U.S. Department of Labor statistics.^23^

#### PSSRQ survey

The quality of the provider was assessed using parental responses to the PRSSQ. Average scores from the component surveys of the PRSSQ were summed and given weight to adjust for their differing sizes of contributions to the aggregated PRSSQ metric.

One month after completion of patient treatment, the PRSSQ was administered to parents or legal caregivers of the patients via telephone or in-person.

#### Bias

The primary sources of bias were selection and measurement error. Selection bias was addressed by recruiting a large sample of subjects across a large and diverse population in seven states. Measurement errors were addressed by orienting and training the data collectors on how to measure the direct labor costs and how to objectively collect PRSSQ survey data for each subject.

#### Statistical Methods and Quantitative Variables

Means and standard deviations were calculated for continuous variables. Counts and percentages were calculated for categorical variables. These descriptive statistics were presented both overall and by treatment modalities. Comparisons of distributions by treatment modalities were conducted by the Chi-square test (or the Fisher’s exact test, as appropriate) for categorical variables and by the one-way ANOVA test (or the Kruskal-Wallis test, as appropriate) for continuous variables.

To compare the value score by states and by facilities in which patients received their treatment, Tukey’s multiple comparison test was used. Preliminary data analysis demonstrated there was significant difference in the value score among three treatment modalities. Hence, comparisons of the value score by states and by facilities were conducted within each category of treatment modality. Natural logarithm transformation was applied to the value score to approximate a normal distribution. States or facilities with less than ten observations were excluded from the multiple comparison. Letter grade ratings were assigned to provider groups which were clustered to have statistically similar mean value scores.

To assess the robustness of results based on the Tukey’s multiple comparison test to potential heterogeneity of variances of the ln-transformed value score across states and facilities, a sensitivity analysis using the non-parametric Dunn’s Kruskal-Wallis multiple comparison test was conducted.

All analyses were conducted in R version 3.5.0 for Windows,^24^ with ‘lsmeans’ package for Tukey’s multiple comparison test,^25^ and ‘FSA’ package for Dunn’s Kruskal-Wallis multiple comparison test.^26^

## RESULTS

### Participants and Descriptive Data

The analytic sample population included 487 patients, with an average age of 3.99 years (standard deviation (**sd**)=1.22) and characteristics comprised predominantly of female (50.1 percent), Hispanic ethnicity (56.2 percent), Other race (45.2 percent), and Medicaid enrollees (94.9 percent). The distribution by treatment modality group was SCGA (53 percent), SC (32 percent), and NSC (15 percent) (Table 1). Significant differences in the distributions of nearly all demographic variables were observed among three treatment modalities. Significant differences were also observed in the quality, cost, and value score. Specifically, NSC had on average the highest quality score, lowest cost score, and hence the highest value score among the three treatment modalities. The rating hierarchy of the value score was NSC > SC > SCGA. Comparisons of the value score by states and facilities were therefore carried out within each treatment modality.

**Table 1.**
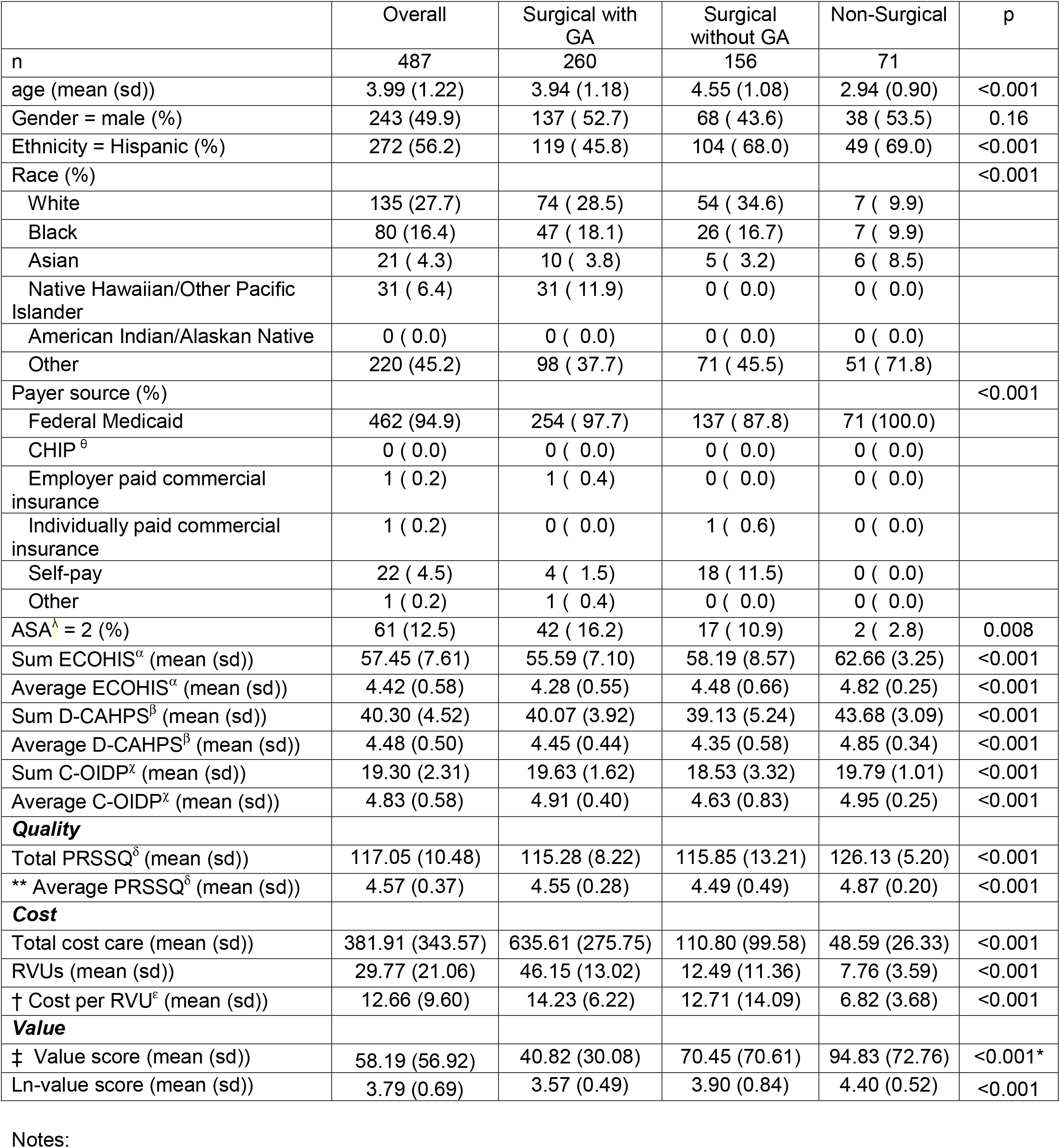
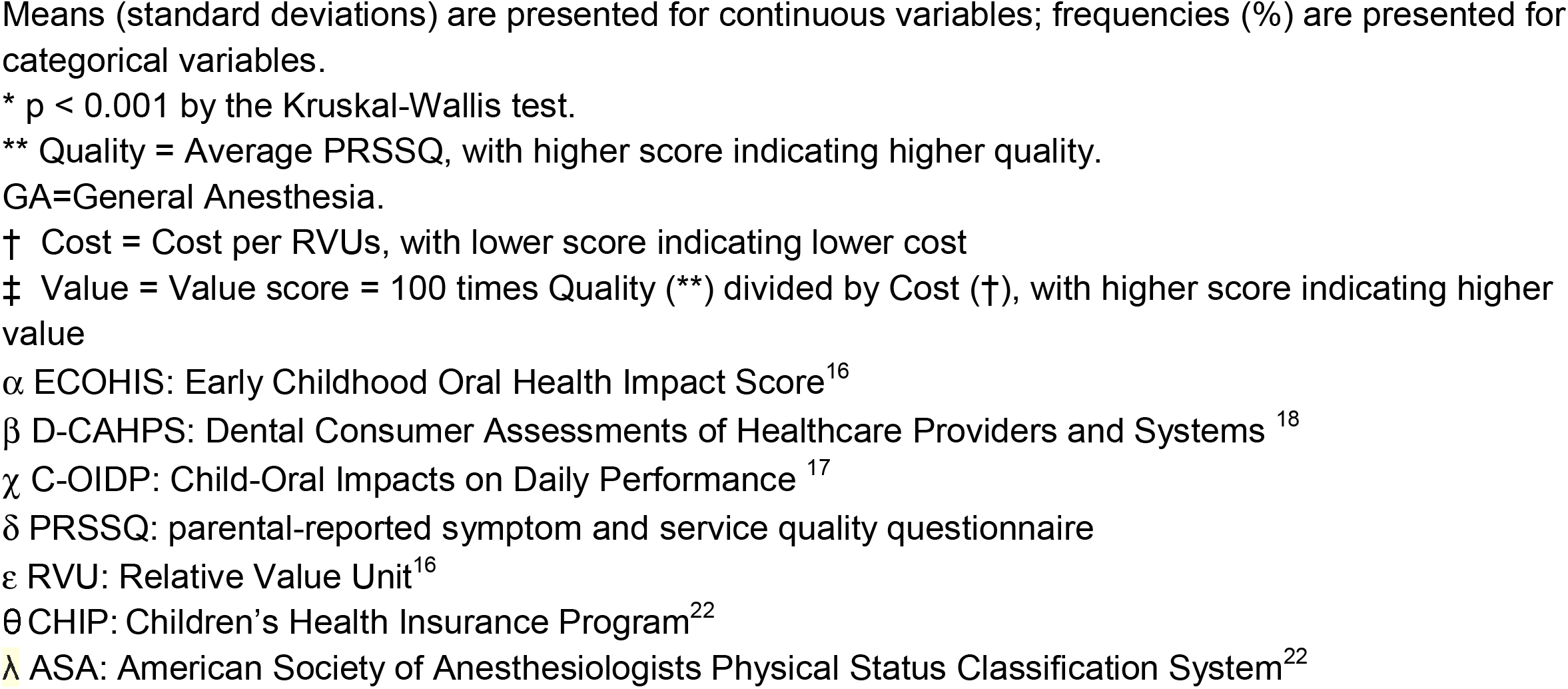
Summary of sample characteristics and comparisons by treatment modalities

### Outcomes Data and Main Results

Figure 2 presents the results from Tukey’s multiple comparison test for the comparisons of the ln-transformed value score by states, with all results significant at p<0.001. For SCGA, Tennessee had the highest average value score with letter grade (A), followed by the respective states and letter grades of Arizona (A-), New York (B), and Hawaii (B-). These four states also have on average significantly higher value scores than the other three states with letter grades of (C), namely, Florida, Maryland, and Massachusetts, with no significant differences observed between these three states. For SC, Maryland had significantly higher value score and letter grade (A) than the other states, followed by Arizona (B), New York (B-), Tennessee (C). The value score for these four states outnumbered that of Florida and Massachusetts, both with letter grade (D). For NSC, patients were only seen in Maryland and New York. The respective (letter grade) and [ln-value mean score (standard deviation)] were significantly higher for Maryland (A) [4.81 (0.60)] than that of New York (B) [4.30 (0.44)].

**Figure 2.**
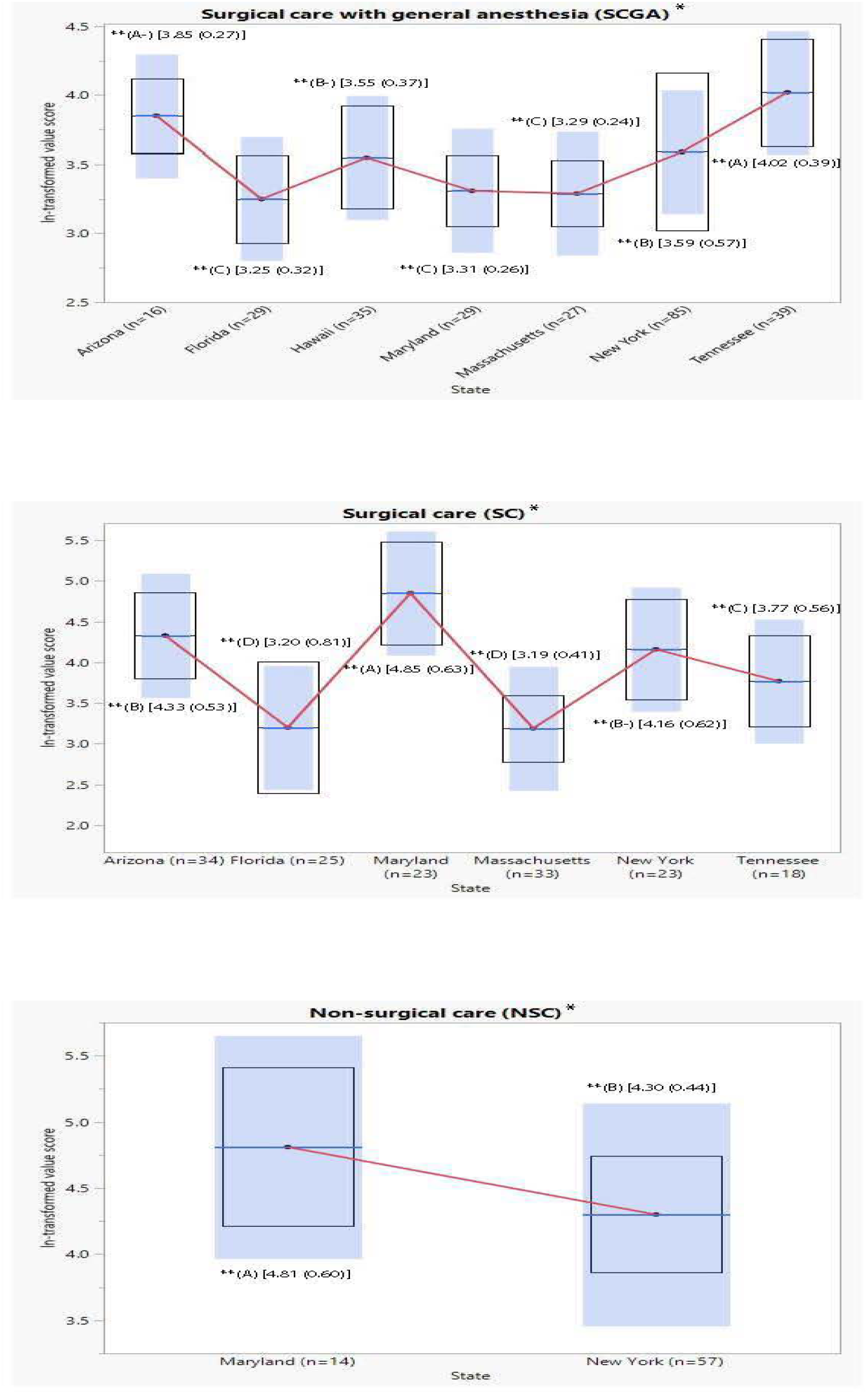
Boxplots of ln-transformed value score by state. Tukey’s multiple comparison test was used. *Treatment modality group **(Letter grade) [mean ln-value score (standard deviation)] GA=General Anesthesia. A(-) is not significantly different than A or B. B(-) is not significantly different than B or C.

Figure 3 shows the results from Tukey’s multiple comparison test by facilities, with all results significant at p<0.001. For SCGA, facilities with letter grade (A) included TN-2 and AZ-1, followed by letter grade (B) at HI-1, FL-4, NY-2, MA-2, and MD-2. For SC, the letter grade groups included (A) at MD1; (B) at AZ-1; (B-) at NY-1; (C-) at TN-1; (D-) at FL-2; and (E) at MA- For NSC, patients were only seen at MD-1 and NY-1. The respective (letter grade) and [ln-value mean score (standard deviation)] were significantly higher for MD-1 (A) [4.81 (0.60)] than for NY-1 (B) [4.30 (0.44)].

**Figure 3.**
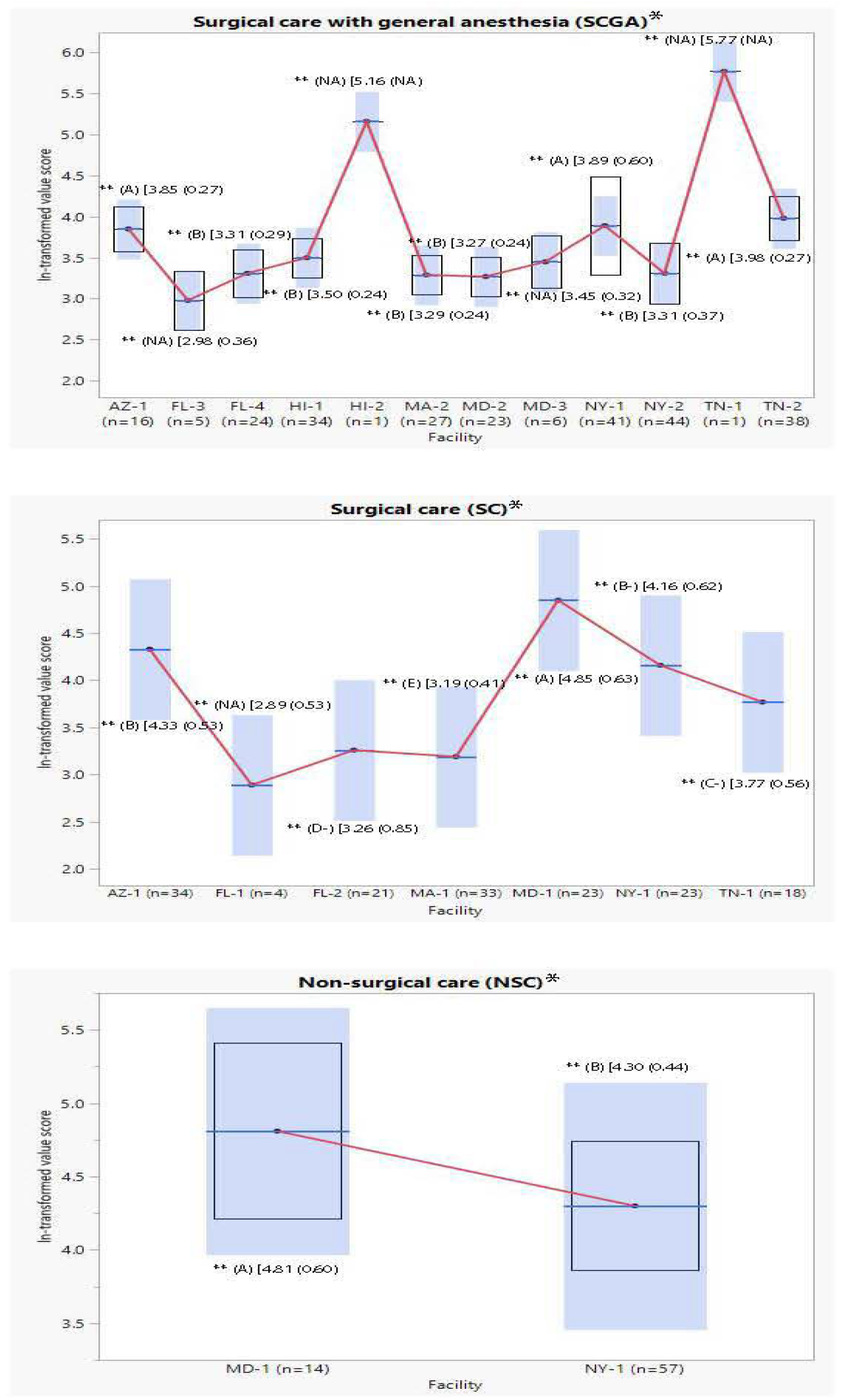
Boxplots of ln-transformed value score by facilities. Tukey’s multiple comparison test was used. *Treatment modality group **(Letter grade) [mean ln-value score (standard deviation)] GA=General Anesthesia. B(-) is not significantly different than B or C. C(-) is not significantly different than C or D. D(-) is not significantly different than D or E

To assess the robustness of results based on the Tukey’s multiple comparison test to potential heterogeneity of variances of the ln-transformed value score across states and facilities, a sensitivity analysis using the non-parametric Dunn’s Kruskal-Wallis multiple comparison test was conducted. Table 2 presents the results of the sensitivity analysis by state and Table 3 shows the results by facility. The sensitivity analyses based on the Dunn’s Kruskal-Wallis multiple comparison test yielded similar results and confirmed the findings shown in Figures 2 and 3.

**Table 2.** Comparisons of value scores (median [IQR]) by states. Sensitivity analysis based on the Dunn’s Kruskal-Wallis multiple comparison test.

| State | Surgical with GA |  |  | Surgical without GA |  |  | Non-surgical |  |  |
| --- | --- | --- | --- | --- | --- | --- | --- | --- | --- |
|  | N | Value (median [IQR]) | Letter Grade Rating* | N | Value (median [IQR]) | Letter Grade Rating * | N | Value (median [IQR]) | Letter Grade Rating* |
| Arizona | 16 | 48.5<br>[37.6, 54.6] | A | 34 | 66.86<br>[50.45, 113.34] | A(-)<sup>α</sup> | - | - |  |
| Hawaii | 35 | 33.6<br>[28.9, 40.0] | B | - | - | - | - | - |  |
| Florida | 29 | 26.2<br>[21.7, 31.9] | C | 25 | 23.57<br>[19.99, 50.87] | D(-)<sup>δ</sup> | - | - |  |
| Massachusetts | 27 | 27.2<br>[23.1, 31.6] | C | 33 | 24.20<br>[20.77, 28.49] | E | - | - |  |
| Maryland | 29 | 27.0<br>[22.7, 32.7] | C | 23 | 140.68<br>[78.97, 187.01] | A | 14 | 108.77<br>[86.44, 152.66] | A |
| New York | 85 | 32.8<br>[24.0, 51.4] | B | 23 | 53.92<br>[42.66, 86.95] | B(-)<sup>β</sup> | 57 | 73.11<br>[56.68 101.30] | B |
| Tennessee | 39 | 54.5<br>[47.9, 62.3] | A | 18 | 45.50<br>[26.50, 73.96] | C(-)<sup>χ</sup> | - |  |  |
| p-value* |  | <0.001 |  |  | <0.001 |  |  | 0.004 |  |

IQR: inter-quartile range.
\* overall test difference.
GA=General Anesthesia.
α: A(-) is not significantly different than A or B.
β: B(-) is not significantly different than B or C.
χ: C(-) is not significantly different than C or D.
δ: D(-) is not significantly different than D or E.

**Table 3.** Comparisons of value scores (median [IQR]) by facilities. Sensitivity analysis based on the Dunn’s Kruskal-Wallis multiple comparison test.

| State-Facility Number | Surgical with GA |  |  | Surgical without GA |  |  | Non-surgical |  |  |
| --- | --- | --- | --- | --- | --- | --- | --- | --- | --- |
|  | N | Value (median [IQR]) | Letter Grade Rating* | N | Value (median [IQR]) | Letter Grade Rating* | N | Value (median [IQR]) | Letter Grade Rating* |
| AZ-1 | 16 | 48.53<br>[37.55, 54.55] | A(-)<sup>α</sup> | 34 | 66.86<br>[50.45, 113.34] | A(-)<sup>α</sup> |  |  |  |
| HI-1 | 34 | 32.90<br>[28.85, 38.89] | C | - | - |  |  |  |  |
| HI-2* | 1 | 174.3<br>[174.3, 174.3] | - | - | - |  |  |  |  |
| FL-1 | - | - | - | 2 | 24.34<br>[22.80, 25.87] | - |  |  |  |
| FL-2 | - | - | - | 21 | 24.36<br>[19.99, 51.49] | D(-)<sup>δ</sup> |  |  |  |
| FL-3 | 5 | 21.11<br>[15.58, 22.15] | D | - | - |  |  |  |  |
| FL-4 | 24 | 27.37<br>[24.27, 31.95] | C(-)<sup>χ</sup> | - | - |  |  |  |  |
| MA-1 | - | - | - | 33 | 24.20<br>[20.77, 28.49] | E |  |  |  |
| MA-2 | 27 | 27.19<br>[23.05, 31.55] | C(-)<sup>χ</sup> | - | - |  |  |  |  |
| MD-1 | - | - | - | 23 | 140.68<br>[78.97, 187.01] | A | 14 | 108.77<br>[86.44, 152.68] | A |
| MD-2 | 23 | 26.39<br>[21.96, 31.27] | D | - | - |  |  |  |  |
| MD-3 | 6 | 34.56<br>[28.20, 37.95] | C(+/-)<sup>ε</sup> | - | - |  |  |  |  |
| NY-1 | 41 | 40.44<br>[31.58, 73.21] | A(-)<sup>α</sup> | 23 | 53.92<br>[42.66, 86.95] | B(-)<sup>β</sup> | 57 | 73.11<br>[56.68, 101.30] | B |
| NY-2 | 44 | 26.79<br>[20.51, 33.42] | C(-)<sup>χ</sup> | - | - |  |  |  |  |
| TN-1* | 1 | 320.3<br>[320.3, 320.3] | - | 18 | 45.50<br>[26.50, 73.96] | C(-)<sup>χ</sup> |  |  |  |
| TN-2 | 38 | 54.44<br>[47.49, 61.45] | A | - | - |  |  |  |  |
|  |  | <0.001 |  |  | 0.001 |  |  | 0.004 |  |

\* facilities with n <5 did not enter statistical testing
\*\*overall test of difference.
GA=General Anesthesia.
α: A(-) is not significantly different than A or B.
β: B(-) is not significantly different than B or C.
ε: C(+/-) is not significantly different than B or D
χ: C(-) is not significantly different than C or D.
δ: D(-) is not significantly different than D or E.

## DISCUSSION

### Findings and Hypothesis

The key results showed statistical significance for rating providers by geographic region and specific clinic facilities. For analyzed U.S. states, Arizona and Tennessee received the highest mean ln-value scores with the letter grade (A) rating for SCGA; Maryland received the highest mean ln-value score with letter grade (A) rating for SC and NSC. For analyzed clinic facilities, two dental clinics with mobile dentist-anesthesiology services, AZ-1 and NY-1, and a surgery center, TN-2, received the highest mean ln-value scores with letter grade (A) rating for SCGA; Maryland received the highest mean ln-value score with letter grade (A) for SC and NSC.

This study’s findings rejected the null hypothesis and demonstrated that the statistical model identified distinct geographic and clinic facility provider groups, which were rated by mean ln-value scores, and associated letter grade ratings. This evidence-based model demonstrated a hierarchal, value-based, health care provider rating system.

### Comparison to Literature

Except for another article by this study’s principal investigator,^27^ a search of the literature did not yield studies which quantified oral health care value by quality and cost. Instead, the literature exclusively focused on either the rationale for value-based health care, quality (i.e., patient-reported and/or clinical outcomes), or cost.

Multiple studies support the findings presented here regarding the rationale of value-based rating systems. In 2008 the Institute of Medicine (renamed as the National Academy of Medicine in 2015) and the Institute for Healthcare Improvement developed a framework of the “Triple Aim” goals for the future of high-value U.S. health care delivery, which consisted of “improving the individual experience of care, improving the health of populations, and reducing the per capita costs of care for populations.^28^” A 2021 white paper from the University of Pennsylvania Leonard Davis Institute (**LDI**) of Health Economics looked at how far the U.S. health care system has progressed toward establishing a value-based institution since the passage of the Affordable Care Act (**ACA**) in 2010 and concluded that reimbursement systems should “push providers away from fee-for-service payment” and well-designed alternative payment models have resulted in improved value with cost savings.^29^

Quality-focused publications, which included patient-reported and clinician-outcomes studies, were found in the dental literature. One research team found that online reviews in platforms such as Google, Yelp, and Healthgrades form a large and rich dataset reflective of the patient experience, which can have the effect of creating a de facto patient-reported quality metric for providers.^30^ However, these websites are more susceptible to selection bias depending on the demographic of the patients that choose to leave publicly visible reviews.

A collaboration between the *Fédération Dentaire Internationale* (**FDI**) World Dental Federation and International Consortium for Health Outcomes Measurement (**ICHOM**) developed an Adult Oral Health Standard Set (**AOHSS**) of patient- and clinician-reported outcome measures, specifically focusing on caries and periodontal disease.^31^ One author outlined the challenges for developing value-based oral health care reporting system, which included the need for creating multi-disciplinary care, creating quantitative value measurement, identifying cost attribution for care, and establishing bundled care reimbursement models.^32^

Cost-focused medical research on peri-operative care,^33^ musculo-skeletal and diabetes care,^34^ cystectomy,^35^ and extracorporeal life support^36^ exclusively emphasized the identification, measurement, and implementation of cost efficiencies. One author reported that, although “medicine has built a body of knowledge from cost-effectiveness, cost-benefit and cost-utility studies, dentistry has not addressed the same questions to any significant degree” and, thus, without a firm estimation of cost, an accurate estimate of value cannot be determined.^37^

### Study Strengths, Limitations, and Generalizability

Strengths of this study include geographic diversity, given the range of states that were included. The patient population also had extensive racial and ethnic diversity. Furthermore, the post-hoc power analysis yielded a power level of 99.4 percent.

One of the limitations of this study is that the PRSSQ quality score in this study was a subjective value. Another limitation is that the sample population was recruited at FQHCs, which serve low SES populations where the cost and quality variables may differ compared to non-FQHC populations.

The findings may not be generalizable due to the high percentage of Medicaid enrollees in the study. Additionally, direct labor costs were used as a proxy for total cost, which could skew the cost metric. The study was also limited by the number of observations of the individual providers. Furthermore, there was no randomization process for assigning patients to providers and some of the states did not see patients for each of the three procedure types, limiting the analysis of provider value by facility for a given treatment.

### Future Studies

Future studies might apply this model of value-based health care to incorporate a sufficiently large data set at the individual provider level for greater granularity for the provider rating system. Additionally, a randomized, controlled study design which includes both FQHC and non-FQHC populations would strengthen the validation of the PRSSQ survey and of the value model.

### Significance and Implications

This study allowed for the use of an evidence- and value-based approach to assess provider value in a standardized format. With some potential modifications to allow for this system to be used at a single provider level, these findings will allow for the instrumentation of an evidence-based modality for ranking providers by value, which can inform patients in their decision of where to seek care, motivate providers to increase value, and provide data so payers might identify high-value providers.

## CONCLUSION

Based on the results of this study, the following conclusions can be made:

1. An objective, value- and evidence-based rating system of providers, by geographic region and treatment facility, can assist:
  a. Patients to better compare and select providers.
  b. Providers to benchmark their comparative ratings on an internal and external basis for continuous quality improvement.
  c. Third-party payers, such as insurance companies and government programs, to better compare outcomes and costs for provider systems and clinic facilities.

## Data Availability

All data produced in the present study are available upon reasonable request to the authors.

## Abbreviations in sequential order of first presentation

(U.S.): United States
(CMS): Centers for Medicare and Medicaid Services
(ICHOM): International Consortium for Health Outcomes Measurement
(ADA): American Dental Association
(DQA): Dental Quality Alliance
(ECC): early childhood caries
(FQHC): federally qualified health center
(NSC): minimally invasive non-surgical care
(SC): surgical restorative care without adjunctive sedation or general anesthesia
(SCGA): surgical restorative care with general anesthesia
(PRSSQ): parental-reported symptom and service quality questionnaire
(SDF): silver diamine fluoride
(HCP): health care providers
(GA): general anesthesia
(Q): quality
(ECOHIS): Early Child Oral Health Impact Scale
(C-OIDP): Child-Oral Impacts on Daily Performance
(D-CAHPS): Dental Consumer Assessments of Healthcare Providers and Systems
(C): cost
(RVU): Relative Value Units
(V): value
(ASA): American Society of Anesthesiologists
(LDI): University of Pennsylvania Leonard Davis Institute of Health Economics
(ACA): Affordable Care Act
(FDI): *Fédération Dentaire Internationale*
(AOHSS): Adult Oral Health Standard Set

## ACKNOWLEDGMENTS

The authors extend appreciation and acknowledgments to Jay Balzer, DDS, MPH, a dental public health specialist and reviewer, and the former NYU Langone Hospitals-Advanced Education in Pediatric Dentistry residents who supported the study as research-researchers: Jared Cattron, DMD, Danielle Fernandez, DMD, Shanti Gopalan, DDS, Mehedia Haque, DMD, MPH, Mariam Javaid, DMD, Stephany Liu, DDS, Andrea Lochan, DMD, Jeremy Morris, DDS, Olanrewaju Oye-Somefun, DDS, Andrea Salazar, DDS, Erin Saucier, DMD, Silpa Velivela, DMD, and Pavel Zeylikman, DMD.

